# How comprehensive is the Cochrane Central Register of Controlled Trials for identifying clinical trial registration?: the protocol of a diagnostic study

**DOI:** 10.1101/2019.12.26.19014274

**Authors:** Masahiro Banno, Yasushi Tsujimoto, Yuki Kataoka

## Abstract

**Objectives:** To evaluate whether or not systematic reviewers can use Cochrane Central Register of Controlled Trials (CENTRAL) to identify ongoing and unpublished studies instead of searching International Clinical Trials Registry Platform (ICTRP) and ClinicalTrials.gov (CT.gov).

**Methods:** This will be a diagnostic accuracy test study. We will collect a consecutive sample of ongoing or unpublished studies on Cochrane Database of Systematic Reviews (CDSRs) during the last six months. We will use all of the records as our reference standard and evaluated whether they are part of the CENTRAL search presented in the CDSRs.

The index test is the CENTRAL search using the search terms in the CDSRs, and the reference standard is the list of ongoing or unpublished studies registered on the ICTRP or CT.gov in the CDSRs.

We will assess the sensitivity and number needed to read.

**Ethics & Dissemination:** This study does not require ethics approval. We registered this study protocol. We will publish the findings in a peer-reviewed journal and may present them at conferences.

**Discussion:** This study may lessen the burden of systematic reviewers if this study clarifies CENTRAL search can be used for screening in records about ongoing or unpublished studies.

**Registration:** UMIN-CTR: UMIN000038981

**Author name abbreviations:** Masahiro Banno (MB), Yasushi Tsujimoto (YT), Yuki Kataoka (YK)

## BACKGROUND

Searching for ongoing or unpublished studies when conducting systematic reviews is important to address publication bias [1]. Cochrane handbook and previous studies require systematic reviewers to search both International Clinical Trials Registry Platform (ICTRP) and ClinicalTrials.gov (CT.gov) to identify ongoing or unpublished studies [1–4].

However, searching ICTRP directly is problematic because number of records downloadable per one time is limited and because ICTRP do not return search results with complex search strategies [3, 5].

We can recently search records about ongoing or unpublished randomized controlled trials and quasi-randomized controlled trials on the Cochrane Central Register of Controlled Trials(CENTRAL) [6]. All the records on ICTRP and CT.gov before April 2019 have been included in CENTRAL. Newly identified CT.gov and ICTRP records are added to CENTRAL on a monthly basis [7].

Whether systematic reviewers can search CENTRAL instead of ICTRP and CT.gov to screening records about ongoing or unpublished studies is unknown. If CENTRAL search can replace the search of ICTRP or CT.gov, we expect to identify all ongoing and unpublished studies on ICTRP or CT.gov included in systematic reviews by CENTRAL search alone. The objective of this study is to evaluate the sensitivity and number needed to read (NNR) of CENTRAL search to identify ongoing or unpublished studies for systematic review production.

## MATERIALS AND METHODS

This publication is the full study protocol. This protocol has been registered in the University Hospital Medical Information Network Clinical Trials Registry (UMIN-CTR). The registration number is UMIN000038981.

### Study design

This study is a diagnostic test accuracy study. We will collect a consecutive sample of ongoing or unpublished records on Cochrane Database of Systematic Reviews (CDSRs). We will use the records as our reference standard and evaluate whether they are part of the CENTRAL search. Figure 1 and Figure 2 summarize the concept and design of this study. We define CDSR records as ongoing or unpublished studies including each CDSR. We define CENTRAL records as records derived from CENTRAL search. We define CDSR/CENTRAL records as ongoing or unpublished studies including each CDSR and records derived from CENTRAL search.

**Figure 1:**
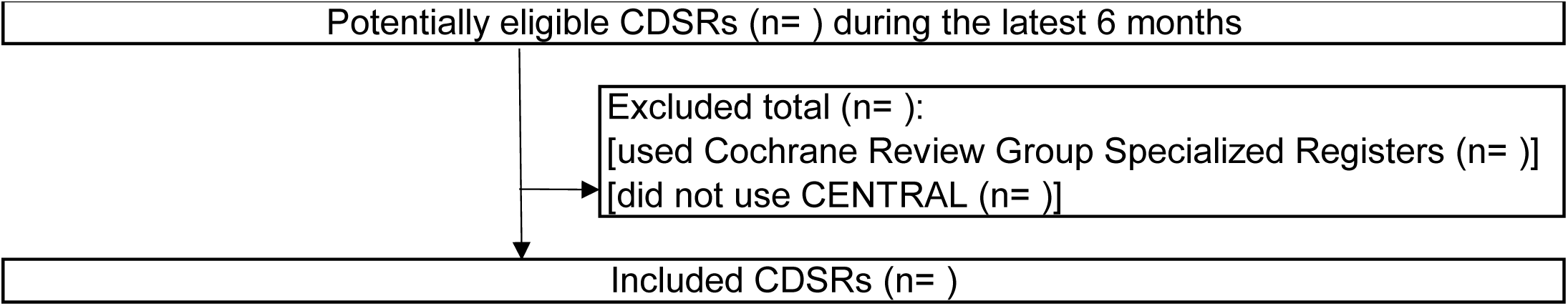
Diagram about Cochrane Database of Systematic Reviews (CDSRs) Abbreviations: CDSRs, Cochrane Database of Systematic Reviews; CENTRAL, Cochrane Central Register of Controlled Trials.

**Figure 2:**
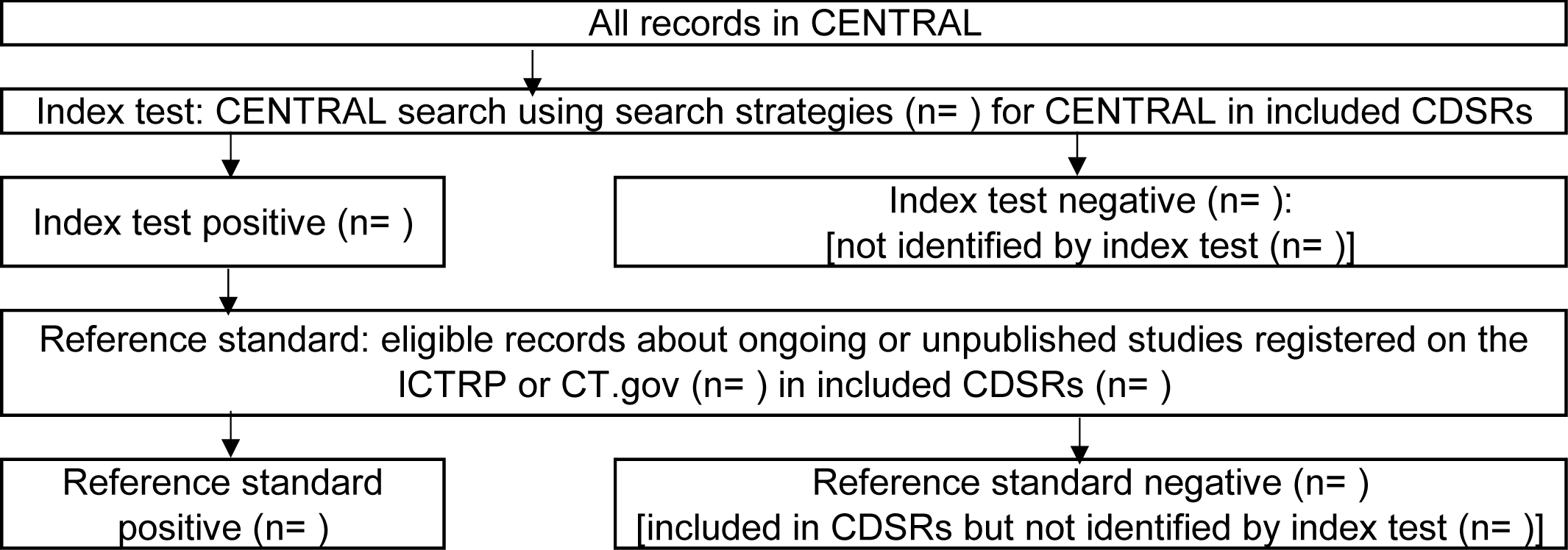
Diagram about registration records Abbreviations: CDSRs, Cochrane Database of Systematic Reviews; CENTRAL, Cochrane Central Register of Controlled Trials; CT.gov, ClinicalTrials.gov; ICTRP, International Clinical Trials Registry Platform.

### Eligible criteria

We will include all records about ongoing or unpublished studies registered on the ICTRP or CT.gov in the latest interventional CDSRs. We will include CDSRs during the last six months. We will exclude CDSRs that used Cochrane Review Group Specialized Registers or CDSRs that did not use CENTRAL, ICTRP, or CT.gov.

To evaluate whether the records of ongoing and unpublished studies in CDSRs can be identified by CENTRAL search alone, we will also include all records of the ICTRP or CT.gov that are identified by the search strategy of CENTRAL presented in CDSRs.

For eligible records, we will extract the following characteristics; trial identifying number, principal investigator, and year of registration.

### Index test and reference standard

The index test will be CENTRAL search. We will manually search CENTRAL with the search strategy presented in each CDSR, in limited publication date, which corresponds to the search date in each CDSR.

The reference standard is the list of ongoing or unpublished studies registered on the ICTRP or CT.gov in the CDSRs, and two other authors will confirm the records. We will tackle disagreements by discussion between the authors. We will choose CDSRs as the data source because CDSRs are performed by rigorous methods following Cochrane Handbook and expected to be available sufficient search strategy to perform a comprehensive CENTRAL search [1].

### Sample size

We do not calculate suitable sample size because this study is an explanatory study. We will focus on CDSRs during the last six months to obtain a certain number of records.

### Data analysis

Our primary outcome is a sensitivity, with its 95 % confidence interval, of CENTRAL search to identify all ongoing and unpublished records. We will calculate a sensitivity, dividing the number of CDSR/CENTRAL records by the number of CDSR records [1]. We will calculate 95 % CI in Wilson score interval with continuity correction by a calculator [8]. We will use Wilson score interval with continuity correction because this method is the accurate method than conventional method [9].

The secondary outcome is a NNR. NNR is a measure of how many records in a database have to be read to identify one of adequate clinical quality and relevance [10]. We will calculate a NNR, dividing the number of CENTRAL records by the number of CDSR/CENTRAL records [11]. We will also report the numbers of records, which are included CDSRs but are not identified by CENTRAL search.

If the sensitivity was not 100%, we will conduct pre-specified subgroup analysis about the primary outcome as follows: 1) type of intervention on CDSRs (pharmacological intervention or non-pharmacological intervention), 2) version of CDSRs (the first version or updated version).

All statistical analyses will be executed in Stata V.15.1 (StataCorp LLC, College Station, Texas, United States of America) [11].

### Ethics

Ethics approval will not be essential because this study conduct research on research.

## DISCUSSION

This is the first study to investigate the appropriateness whether CENTRAL search instead of ICTRP or CT.gov is sufficient to identify ongoing or unpublished clinical trial registration. Therefore, this study will potentially be able to lessen the burden of systematic reviewers if searching ICTRP or CT.gov for identifying ongoing or unpublished studies is not mandatory based on the results of this study.

This study had some expected limitations. First, not all records which undergo CENTRAL search may be assessed by hand search on CDSRs, the reference standard. The potential reason is that the time lags about records from registration on ICTRP or CT.gov to adding to CENTRAL, which may prevent searching CENTRAL from detecting comprehensive data about ongoing or unpublished studies. We may underestimate sensitivity if this limitation arises (for example, specify number of CDSR records is A, number of CDSR/CENTRAL records is B, number of records added after time lag is X. A is larger than B. Superficial sensitivity=B/A, true sensitivity=(B+X)/(A+X). Superficial sensitivity is smaller than true sensitivity because A>B).

In conclusion, this study will perform an investigation about records about ongoing or unpublished clinical trials. The expected results will clarify whether systematic reviewers can search only CENTRAL to identify ongoing or unpublished studies.

## Data Availability

We have no additional data.

## ACKNOWLEDGEMENTS

We would like to thank the Cochrane Library for managing the Cochrane Central Register of Controlled Trials (CENTRAL).

## CONTRIBUTORS

MB, YT and YK contributed to the conception and design of the research. MB is fully responsible for writing the protocol. All authors gave final approval of the protocol before submission. After the publication of the protocol, we plan for the following contributions by each author: MB will screen CDSR records and extract data. YT and YK will validate the records and data. MB will conduct the data analysis. MB, YT and YK will write the manuscript.

## Provenance and peer review

Not peer reviewed.

## Patient consent for publication

Not required.

## Data Availability Statement

We have no additional data.

